# Automated ICD-O-3 Coding of Real-World Pathology Reports Using Self-Hosted Large Language Models

**DOI:** 10.1101/2025.07.23.25332040

**Authors:** Kamyar Arzideh, René Hosch, Amin Turki, Bahadir Erilymaz, Mikel Bahn, Henning Schäfer, Ahmad Idrissi-Yaghir, Sameh Khattab, Amin Dada, Hideo A. Baba, Dirk Schadendorf, Martin Schuler, Jens Kleesiek, Sylvia Hartmann, Felix Nensa, Julius Keyl

## Abstract

While large language models (LLMs) have shown promise in medical text processing, their real-world application in self-hosted clinical settings remains underexplored. Here, we evaluated five state-of-the-art self-hosted LLMs for automated assignment of International Classification of Diseases for Oncology (ICD-O-3) codes using 21,364 real-world pathology reports from a large German hospital. For exact topography code prediction, *Qwen3-235B-A22B* achieved the highest performance (micro-average F1: 71.6%), while *Llama-3*.*3-70B-Instruct* performed best score at predicting the first three characters (micro-average F1: 84.6%). For morphology codes, *DeepSeek-R1-Distill-Llama-70B* outperformed other models (exact micro-average F1: 34.7%; first three characters micro-average F1: 77.8%). Large disparities between micro- and macro-average F1-scores indicated poor generalization to rare conditions. Although LLMs demonstrate promising capabilities as support systems for expert-guided pathology coding, their performance is not yet sufficient for fully automated, unsupervised use in routine clinical workflows.

## Introduction

The increasing volume of pathology reports in modern healthcare systems has created a pressing need for efficient data extraction using established and emerging standardization methods^1−3^. At the core of this standardization is the International Classification of Diseases for Oncology (ICD-O). Its current version, ICD-O-3, builds on a sophisticated dual classification system including both topographical and morphological codes. Topography codes (C00-C80) identify the anatomical site of the tumor using a letter followed by two digits, with an optional decimal for greater specificity. Morphology codes consist of an “M” prefix followed by five digits (M8000/0-M9989/3) that describe the tumor’s histological characteristics. A complete ICD-O-3 code, such as C50.4 M8500/3 represents an infiltrating ductal carcinoma (/3) of the outer quadrant of the breast, while C34.1 M8070/2 indicates a squamous cell carcinoma in situ (/2) in the upper lobe of the lung. This comprehensive coding system enables detailed classification of cancers, which is essential for epidemiological surveillance in cancer registries, interdisciplinary treatment and research. However, its precision comes at the cost of additional documentation burden for clinical staff, especially if these ICD-O-3 codes are not systematically captured in the electronic health record. This can be for instance the case for national or regional cancer registries, which are installed in most European countries to document the real-world incidence of cancer diagnoses in a given population. Manually scanning pathology reports is a daily routine for hospital personnel in many hospitals^4,5^ and cancer registries. Trained personnel spend extensive hours reviewing and coding reports, making the process not only time-consuming but also susceptible to human error^6^. This vulnerability becomes particularly apparent when dealing with complex cases that require correct interpretation of medical terminology^7^. The resource-intensive nature of manual coding, requiring specialized knowledge and continuous training, along with the growing volume of pathology reports, makes the current approach increasingly unsustainable^8,9^.

Large Language Models (LLMs) have been successfully used in the medical domain, with adaptations ranging from clinical decision support systems^10,11^ to assistance for medical documentation^12,13^. LLMs have demonstrated remarkable capabilities in understanding and processing medical text, including pathology reports^14,15^. Despite these promising results, several research gaps remain. First, existing studies often focus on LLMs’ ability to generate or replicate medical language^12,16–18^ rather than specific extraction tasks. Second, existing medical data extraction studies using unstructured reports focus on specific and narrow tasks that do not require extensive reasoning capabilities^19–21^. Third, many publications rely exclusively on large proprietary models like ChatGPT or Gemini to demonstrate clinical applicability.^22–27^ Due to data privacy concerns, however, these models are not suitable for hospital environments. Hence, specific, smaller open-source models must be self-hosted and securely integrated into clinical workflows. Compared to proprietary models such as ChatGPT, open-source models often show lower performance^28–30^ with promising recent improvements^31–33^ introduced by new state-of-the art LLMs like Deepseek-R1 or Qwen-3. Against this background, this study investigates the potential of using state-of-the-art open-source LLMs for ICD-O-3 code extraction from real-world pathology reports at a large comprehensive cancer center.

## Methods

For a visual representation of the methods employed in the study, **Figure 1** illustrates the steps undertaken. These steps are discussed in greater detail in the subsequent subsections.

**Figure 1.**
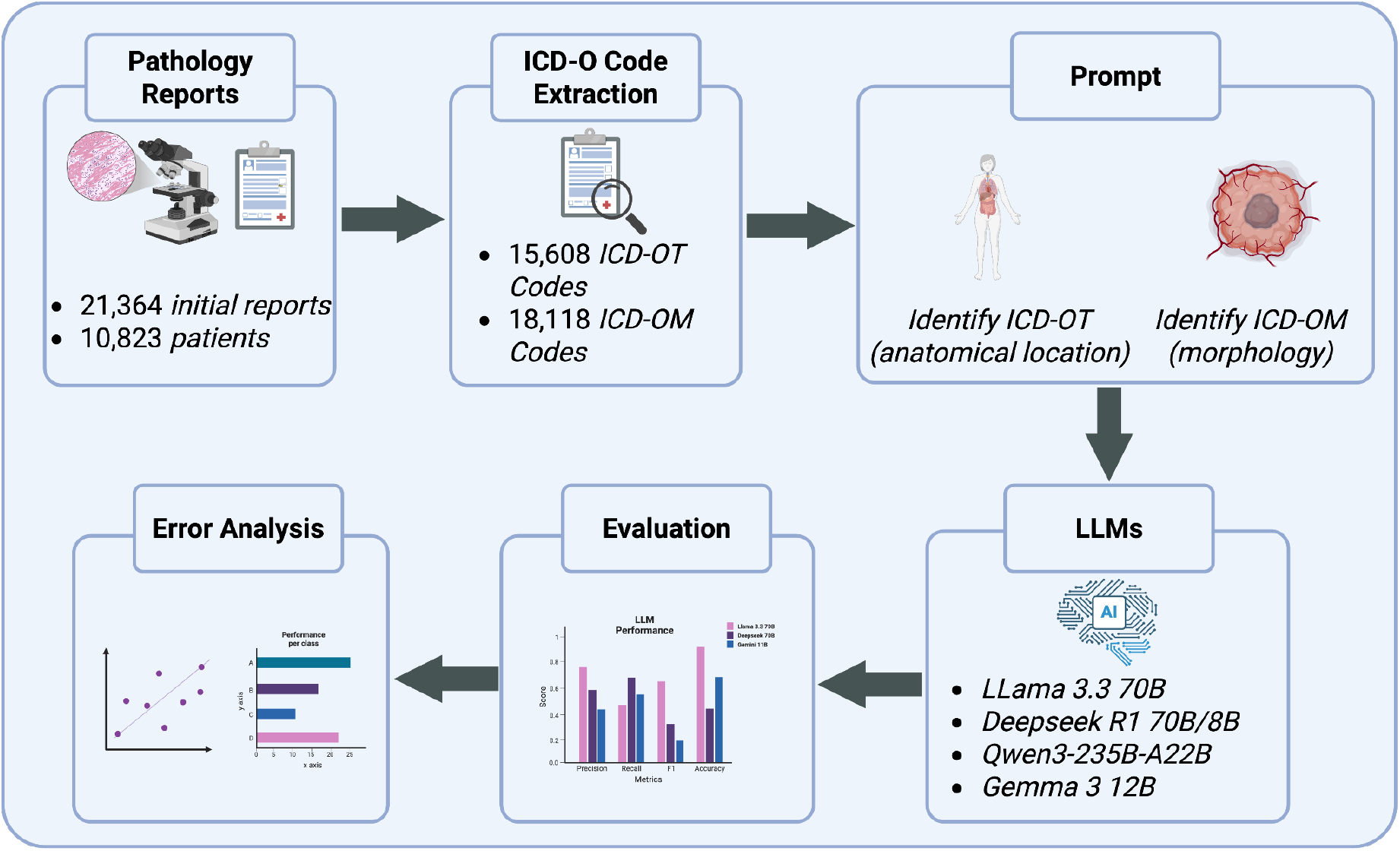
Visual Representation of the Methods employed in the Study to extract ICD-O Codes from Pathology Reports.

### Ethics Statement

This study was approved by the Ethics Committee of the Medical Faculty of the University of Duisburg-Essen (approval number 23-11557-BO). Due to the study’s retrospective nature, the requirement of written informed consent was waived by the Ethics Committee of the Medical Faculty of the University of Duisburg-Essen. All methods were carried out in accordance with relevant guidelines and regulations.

### Dataset

For the study, pathology reports across specialized medical departments, stored in the electronic health record system of the University Hospital Essen, were analyzed. In order to identify reports that contain ICD-O code, the reports were systematically searched using regular expressions. As a result, 21,364 pathology reports from 10,823 patients, documented between 2013 and 2025 during routine hospital operations, were retrieved. These reports contained detailed descriptions of tissue examinations, including anatomical locations and tumor morphology information. In the reports, the ICD-O Topography code (ICD-OT) identifying the anatomical location of the tumor and the ICD-O Morphology (ICD-OM) describing its biological behaviour were manually documented by pathologists. These codes were extracted to form a ground truth and removed from the report so that the model can generate codes without having the correct answer in the input.

ICD-O codes used as ground truth were generated through standard institutional coding procedures following established guidelines^34–36^. These procedures typically adhere to WHO/IARC ICD-O-3 coding standards^34^, German cancer registry requirements^35^, and international best practices for oncology coding^36^.

### Prompt Engineering

We developed a prompt engineering approach to extract relevant information from the pathology reports by LLMs. In total, three different prompts were created and used to extract ICD-O codes from unstructured reports.

The first prompt includes a list of ICD-OT codes together with the corresponding anatomical locations. The LLM was tasked to identify the ICD-OT codes based on the patient-specific pathology reports. The additional context provides additional information about anatomical locations and should help the model with identifying the correct topography codes.

In the second prompt, no additional information about the anatomical locations was provided. The rest of the prompt was identical. This way, the impact of the additional context was evaluated to give insights into the performance differences when providing additional context.

For the third prompt, the morphology of the tumor was asked and the answer should be provided by returning the ICD-OM codes. General information about the ICD-OM codes and how they are structured are provided to help the model to identify the appropriate codes.

The exact formulation of all prompts including an example anonymized pathology report and the answers from the LLM are provided in Supplementary Figure 1-3. To make it easier to evaluate the outputs, all models were prompted to return only the codes without any explanations.

#### Models

In this study, five different open-source models were evaluated. All models were deployed on secure hospital infrastructure to ensure data privacy.

1. *Llama-3*.*3-70B-Instruct*^37^, an open-source model that was optimized for dialogue use cases like question-answering by instruction fine-tuning. The model was pre-trained on multilingual datasets and uses reinforcement learning with human feedback to align with human preferences^37^.
2. *DeepSeek-R1-Distill-Llama-70B*^38^ is a fine-tuned variant of the *Llama-3*.*3-70B-Instruct* LLM and a technique called model distillation was used to enhance the performance. Distillation denotes the process of transferring knowledge from a larger model to a smaller one. This is a cost-effective approach, as smaller models can be employed on less powerful hardware. Distillation was performed by fine-tuning the model on generated datasets from the larger *DeepSeek-R1* model, which is a state-of-the-art 671B parameter reasoning model. According to its authors, the larger model *DeepSeek-R1* achieves performance comparable to OpenAI-o1 across math, code, and reasoning tasks^38^.
3. Besides the 70B model, the smaller *DeepSeek-R1-Distill-Llama-8B* model was also evaluated to take insights into the importance of parameter size. This model is a fine-tuned variant of the *Llama-3*.*3-8B-Instruct* and was trained on the same dataset as the larger model.
4. *Qwen3-235B-A22B*^39^, which is a mixture-of-experts model with reasoning capabilities similar to Deepseek-R1. Users can enable or disable reasoning output explaining the thinking process that the LLM used to solve the task. The mixture-of-experts architecture allows for the dynamic activation or deactivation of components based on the input. During inference, only a maximum of 22B of 235B parameters are activated. For this study, the thinking process was enabled.
5. gemma-3-12B-it developed by Google^40^. The model is part of the *gemma-3* family, which include lightweight multimodal models that can process large contexts up to 128k tokens. The model was primarily chosen to provide a benchmark for the lightweight *DeepSeek-R1-Distill-Llama-8B* model. In **Supplementary Table 1**, an overview of the evaluated models, including information about parameter size and architecture is provided. In **Supplementary Table 2**, the used sampling parameters for inference of the LLMs are documented.

#### Evaluation

The prediction of ICD-O codes was measured by defining it as a multi-label classification task. Each pathology report corresponded to one or more ICD-O codes, which the model had to predict. Performance metrics like precision, recall, and F1-score were calculated by counting the number of true positives, false positives, and false negatives. In order to assess the performance of models on common and rare diseases, both micro- and macro-average score calculation methods were applied. Confidence intervals (CI95%) were reported using 1.000 iterations of bootstrapping^41^. Exact code matching is the most straightforward approach to calculate evaluation metrics. For example, if the ground truth ICD-O code is *C44*.*8* and the predicted code is also *C44*.*8*, this is considered a true positive prediction. However, the differentiation of the last digit of an ICD-code can be challenging, even for human expert annotators. For example, the ICD-code *C44*.*8* refers to “*other malignant neoplasms: skin, overlapping several parts”* and the ICD-code *C44*.*9* stands for “*malignant neoplasm of the skin, unspecified”*. In these specific examples, the difference between these codes is subtle and there is no clear indication that reliably differentiates one from the other. Therefore, in addition to evaluation on the exact code match, a second evaluation was performed on only the first three positions of the ICD code. For example, if the code in the ground truth was *C44*.*8* and the predicted code *C44*.*9*, it was considered as a true positive, because the first three positions (*C44*) match. Both evaluation calculation methods were performed to compare the results of the different models on different aspects.

## Results

### Dataset characteristics

We retrospectively collected 21,364 pathology reports containing at least one ICD-OT or ICD-OM code from our hospital database. The majority of pathology reports were derived from skin surgery (C44, 80.15%), followed by lung samples (C34, 5.60%) and breast (C50, 2.36%). Consequently, the most frequent ICD-OM codes were basal cell carcinoma (809, 33.90%), squamous cell carcinoma (807, 21.33%) and melanoma (872, 10.89%) related. The patients had a mean age of 69 years and 44.9% were female. Detailed dataset characteristics are described in **Table 1**.

**Table 1:** Dataset characteristics

|  | Pathology reports (n=21,364) |
| --- | --- |
| Number of patients | 10,823 |
| Age (years), mean (SD) | 69 (SD: 15.56) |
| Sex (female), n (%) | 4,756 (44.9%) |
| Length of text (number of characters) | median: 2760.0<br>average: 3093.5<br>IQR: 1314.2 (Q1: 2268.0, Q3: 3582.2)<br>min: 870<br>max: 16158 |
| Top 10 ICD-OT Groups | C44 (skin) (n=22,818) (80.15%)<br>C34 (lungs) (n=1,594) (5.60%)<br>C50 (breast) (n=671) (2.36%)<br>C49 (connective and other soft tissue) (n=545) (1.91%)<br>C80 (without localization) (n=337) (1.18%)<br>C22 (liver and intrahepatic bile ducts) (n=262) (0.92%)<br>C18 (large intestine) (n=225) (0.79%)<br>C41 (bone and articular cartilage) (n=158) (0.55%)<br>C78 (secondary respiratory and digestive organs) (n=155) (0.54%)<br>C67 (urinary bladder) (n=137) (0.48%) |
| Top 10 ICD-OM Groups | 809 (skin) (n=9,628) (33.90%)<br>807 (breast) (n=6,059) (21.33%)<br>872 (melanoma) (n=3,092) (10.89%)<br>808 (squamous cell carcinoma) (n=3,025) (10.65%)<br>814 (adenomas) (n=1,510) (5.32%)<br>850 (breast) (n=627) (2.21%)<br>874 (melanoma) (n=530) (1.87%)<br>824 (carcinoid) (n=389) (1.37%)<br>804 (small and non-small cell carcinoma) (n=350) (1.23%)<br>801 (epithelioma) (n=320) (1.13%) |

### Processing performance

All pathology reports served as input for LLM extraction of ICD-OT and ICD-OM codes. However, extraction success varied across models due to context length limitations and differences in output generation patterns. Some models, such as Deepseek, generate reasoning content within “<think>” HTML tags (see **Supplementary Figure 4** for an example), which consumes output tokens and thereby reduces the available input context for processing reports. These technical constraints resulted in different extraction rates across models and tasks (**Figure 2)**.

**Figure 2.**
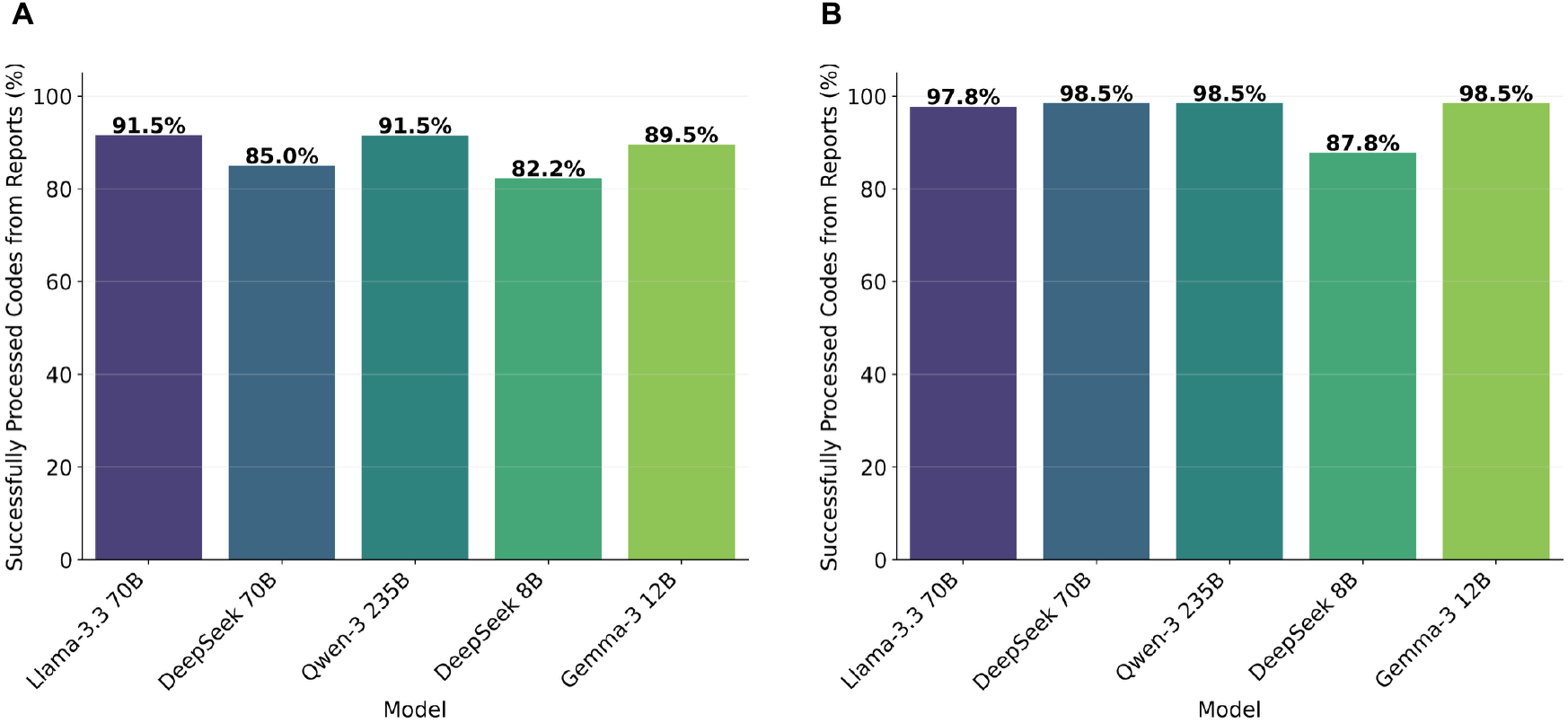
Percentages of ICD-O Codes Extracted from Reports. **A:** ICD-OT Codes, **B:** ICD-OM Codes. The barplots show the percentage of ICD-O codes that could be extracted from pathology reports.

From the initial 21,364 reports, *Llama-3*.*3* could extract 19,554 ICD-OT codes (91.5%, **Fig. 2A**). *Qwen-3* and *Gemma-3* achieved similar results with 19,546 (91.5%) and 19,127 (89.5%) extracted ICD-OT codes. In comparison, the *Deepseek* models only extracted 18,150 (85%) for the *70B* and 17,566 (82.2%) for the *8B* variant. For the ICD-OM task, all models except the *Deepseek-8B* LLM with 18,747 (87.8%) ICD-OM codes could extract a similar number of codes (**Fig. 2B)**.

To ensure fair comparison across models, evaluation was restricted to ICD-O and report pairs where all models generated predictions. This approach prevents bias that would arise from models having different coverage of the dataset, as each model’s performance would otherwise be assessed on a different subset of cases. Only reports for which every model generated an ICD-O code prediction were included in the analysis, yielding 15,608 pairs for the ICD-OT task and 18,118 pairs for the ICD-OM task.

### ICD-O Topography Results

In the exact evaluation setting for extracting ICD-OT codes, where additional information about the anatomical localizations was provided as context, the Qwen3-235B-A22B model achieved the best results with a macro-average F1 score of 29.9% (CI95%: 29.3%-33.1%, **Fig. 3A**) and a micro-average F1 score of 71.6% (CI95%: 70.9%-72.3%, **Suppl. Fig. 5A**). The DeepSeek-R1-Distill-Llama-70B model was the second best performing model with a macro-average F1-score of 26.4% (CI95%: 26.0%-29.5%) and micro-average of 64.3% (CI95%: 63.6%-65.1%). The smaller models were not on par and achieved lower scores, with the DeepSeek-R1-Distill-Llama-8B achieving a micro-average F1-score of 25.4% (CI95%: 24.7%-26.0%) and Gemma-3-12B-it of 19.7% (CI95%: 19.1%-20.3%).

**Figure 3.**
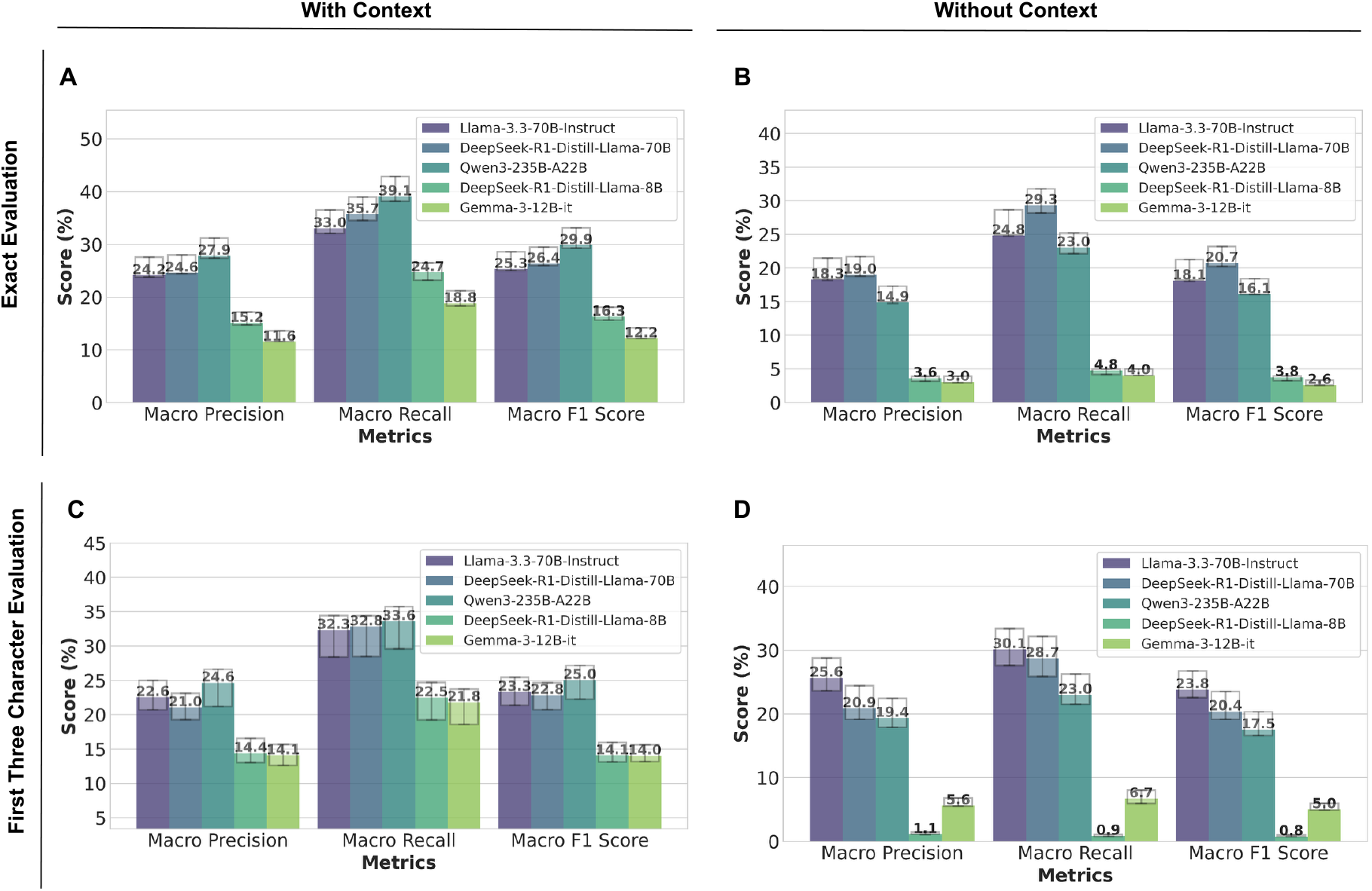
Macro-Average Results of ICD-OT Code Extraction. **A:** Exact Evaluation with Context, **B:** Exact Evaluation without Context, **C:** First Three Character Evaluation with Context, **D:** First Three Character Evaluation without Context. Displayed are results of the different models on the evaluation dataset consisting of 15,608 pairs of pathology reports and ICD-OT codes.

For the results of the evaluation without additional context about the anatomical locations, the *DeepSeek-R1-Distill-Llama-70B* achieved the best results with a macro-average F1-score of 20.7% (CI95%: 20.2%-23.2%, **Fig. 3B**) and micro-average of 42.8% (CI95%: 42.1%-43.6%, **Suppl. Fig. 5B**). *Qwen3-235B-A22B* performed second best with a macro-average F1-score of 16.1% (CI95%: 16.1%-18.4%) and micro-average of 40.4% (CI95%: 39.6%-41.1%). The smaller models again achieved lower scores with the *DeepSeek-R1-Distill-Llama-8B* model achieving a micro-average F1-score 4.3% (CI95%: 4.0%-4.6%) and the *Gemma-3-12B-it* of 8.0% (CI95%: 7.5%-8.4%).

For the first three ICD-code positions with context, the *Llama-3*.*3-70B-Instruct* LLM achieved the best micro-average F1-score of 84.6% (CI95%: 84.0%-85.2%, **Suppl. Fig. 5C**). *Qwen3-235B-A22B* scored second best with a micro-average F1-score of 81.9% (CI95%: 81.3%-82.5%). It also showed the best performance for the macro-average metrics with a F1-score of 25.0% (CI95%: 22.3%-27.1%, **Fig. 3C**). *DeepSeek-R1-Distill-Llama-70B* reached a micro-average F1-score of 80.7% (CI95%: 80.1%-81.3%) and *DeepSeek-R1-Distill-Llama-8B* a micro-average F1-score of 53.6% (CI95%: 52.8%-54.4%).

The evaluation without context showed that the best performing model for predicting the first three characters of ICD-OT codes was also *Llama-3*.*3-70B-Instruct* with a macro average F1-score of 23.8% (CI95%: 22.6%-26.7%, **Fig. 3D**) and a micro-average F1-score of 87.2% (CI95%: 86.6%-87.7%, **Suppl. Fig. 5D**). *DeepSeek-R1-Distill-Llama-70B* scored second best with a macro-average F1-score of 20.4% (CI95%: 19.1%-23.5%) and a macro average F1-score of 81.0% (CI95%: 80.4%-81.6%). Smaller models performed worse with *DeepSeek-R1-Distill-Llama-8B* reaching a micro-average F1-score of 24.2% (CI95%: 23.6%-24.8%) and *Gemma-3-12B-it* of 31.4% (CI95%: 30.7%-32.1%). The detailed results of all experiments are listed in **Supplementary Table 3**.

### ICD-O Morphology Results

Next, we assessed the potential of LLM to extract ICD-O morphology codes from pathology reports. Overall, the *DeepSeek-R1-Distill-Llama-70B* achieved the best results with a macro and micro-average F1-score of 6.5% (CI95%: 6.7%-7.9%, **Fig. 4A**) and 34.7% (CI95%: 34.1%-35.4%, **Suppl. Fig. 6A**) respectively for the exact evaluation. *Qwen3-235B-A22B* was the second best performing model with a macro and micro-average F1-score of 6.0% (CI95%: 6.1%-7.3%) and 22.6% (CI95%: 22.0%-23.2%). *DeepSeek-R1-Distill-Llama-8B* reached a micro-average F1-score of 0.3% (CI95%: 0.2%-0.4%) and *Gemma-3-12B-it* of 7.1% (CI95%: 6.8%-7.5%).

**Figure 4.**
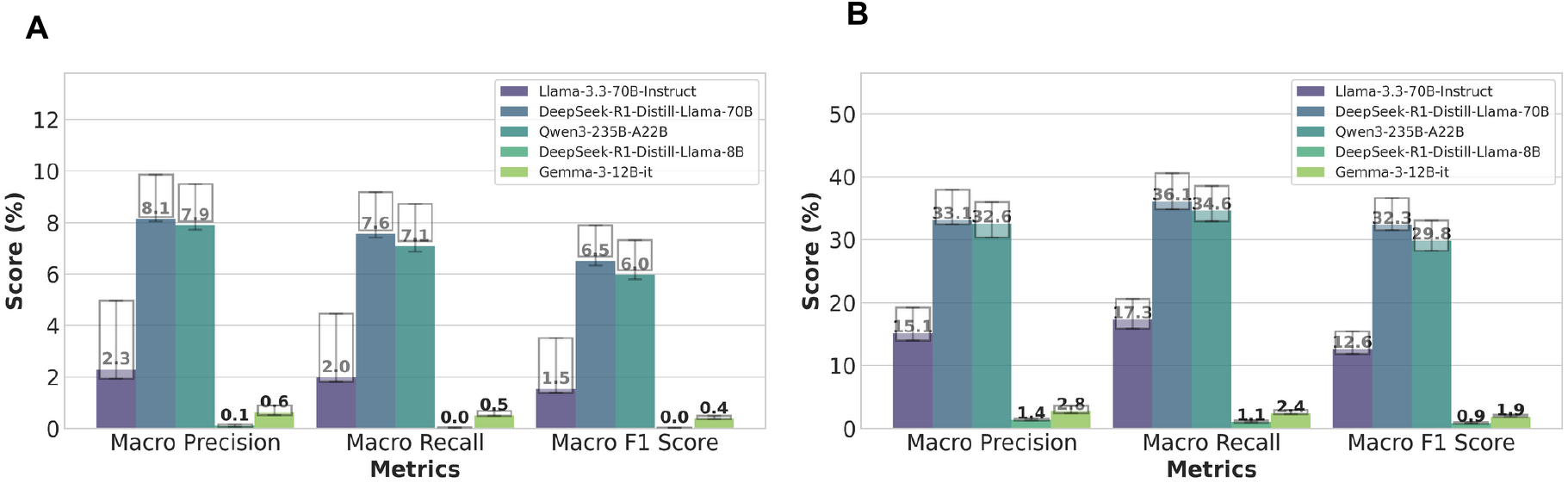
Macro-Average Results of ICD-OM Code Extraction. **A:** Exact Evaluation, **B:** First Three Character Evaluation. Displayed are results of the different models on the evaluation dataset consisting of 18,118 pairs of pathology reports and ICD-OM codes. This evaluation was calculated using exact and first three positions matches of ICD-code prediction.

For the first three character ICD-OM evaluation, *DeepSeek-R1-Distill-Llama-70B* again achieved the highest performance with a macro-average F1-score of 32.3% (CI95%: 31.5%-36.6%, **Fig. 4B**) and a micro-average F1-score of 77.8% (CI95%: 77.2%-78.3%, **Suppl. Fig. 6B**). *Qwen3-235B-A22B* showed the second best results with a F1-macro-average of 29.8% (CI95%: 28.3%-33.1%) and a F1-micro-average of 54.3% (CI95%: 53.6%-55.0%). *DeepSeek-R1-Distill-Llama-8B* and *Gemma-3-12B-it* reached a micro-average F1-score of 4.8% (CI95%: 4.4%-5.1%) and 25.5% (CI95%: 24.9%-26.1%).

### Error Analysis

The evaluation results revealed that LLMs encountered significant challenges in accurately predicting ICD-OT and especially the ICD-OM codes. Differences between micro-and macro-average were small for the ICD-OT task, but larger for the ICD-OM task. This indicated that the performance differed depending on the class. To find out which ICD-classes were difficult for the LLMs to predict, the top five most frequent ICD-OT codes and groups (first three characters) and the corresponding performance of the three best performing LLMs were analyzed.

The exact evaluation scores with context showed high variance in performance between models (**Fig. 5A**). The *Llama-3*.*3-70B-Instruct* model, in particular, performed worse than *DeepSeek-R1-Distill-Llama-70B* and *Qwen3-235B-A22B* for the most frequent classes. Without context, *Llama-3*.*3-70B-Instruct*’s performance dropped drastically (C44.7 decreased from 56.9% to 34.4%, **Fig. 5B**). Overall, ICD-OT codes that appear less frequent in the reports were classified with lower accuracy.

**Figure 5.**
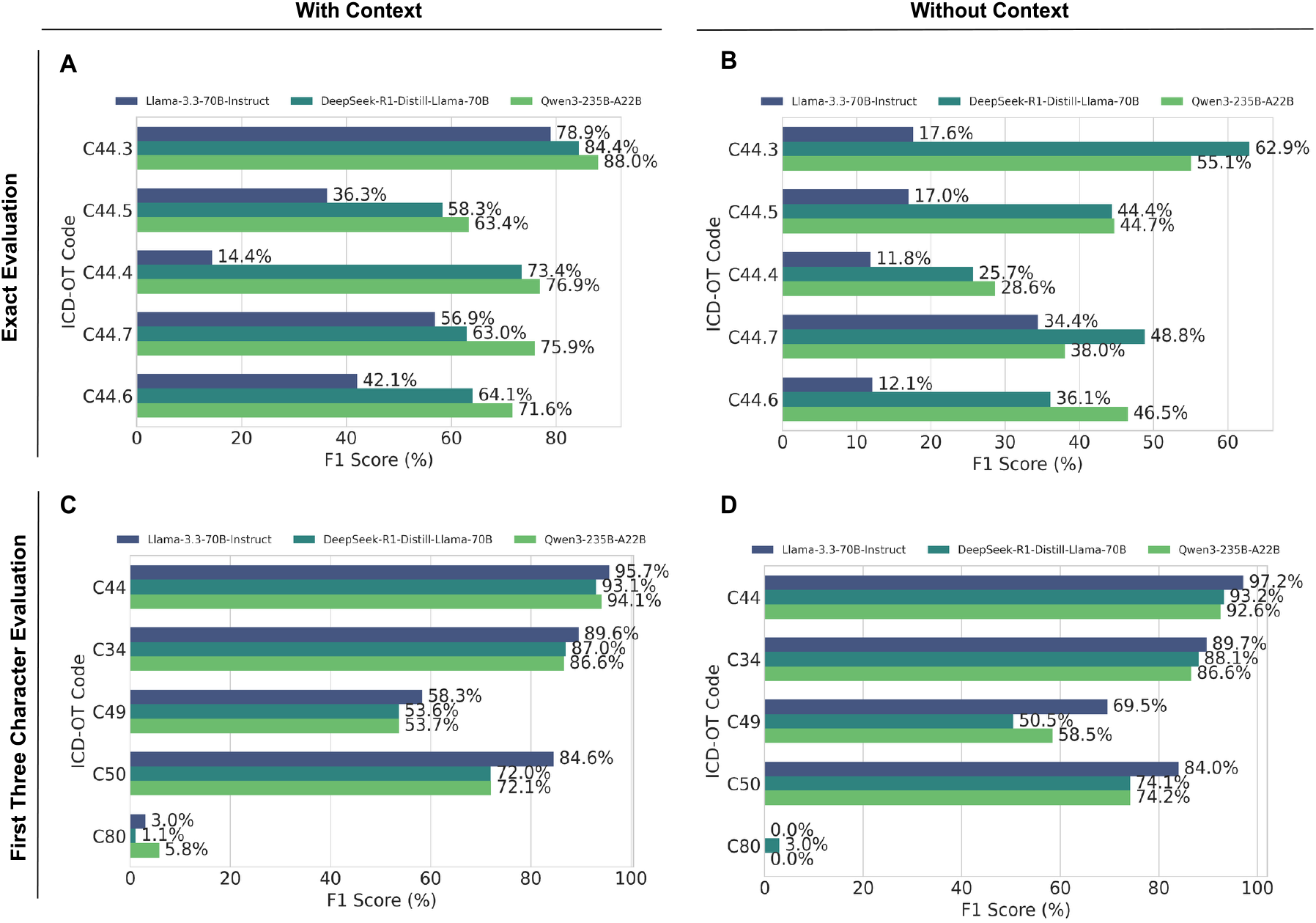
Performance in Relation to Support for ICD-OT Extraction Task. **A:** Exact Evaluation with Context, **B:** Exact Evaluation without Context, **C:** First Three Character Evaluation with Context, **D:** First Three Character Evaluation without Context. Only the performance of the three best models for the five most frequent ICD-OT codes and groups (first three characters) are displayed. The order of the codes is descending, so the code displayed at the top of the plot is the most frequent code.

For the ICD-OT group evaluation with context, the two most frequent ICD-OT groups *C44* and *C43* were extracted with F1-scores ranging between 86.6% and 95.7% (**Fig. 5C**). Other classes with lower support like *C80* showed lower scores varying between 1.1% and 5.8%. For the ICD-OT task without context, the performance per group was similar (**Fig. 5D**). All model performances for each class were largely in the same range, with only minor variation between models.

The exact ICD-OM evaluation showed comparable performance across models (**Fig. 6A**). The most frequent code *8090/3* was extracted with F1-scores ranging from 32% (*Qwen3-235B-A22B*) to 76.3% (*DeepSeek-R1-Distill-Llama-70B*). Other codes, such as *8078/3* or *8081/2*, were classified with lower scores.

**Figure 6.**
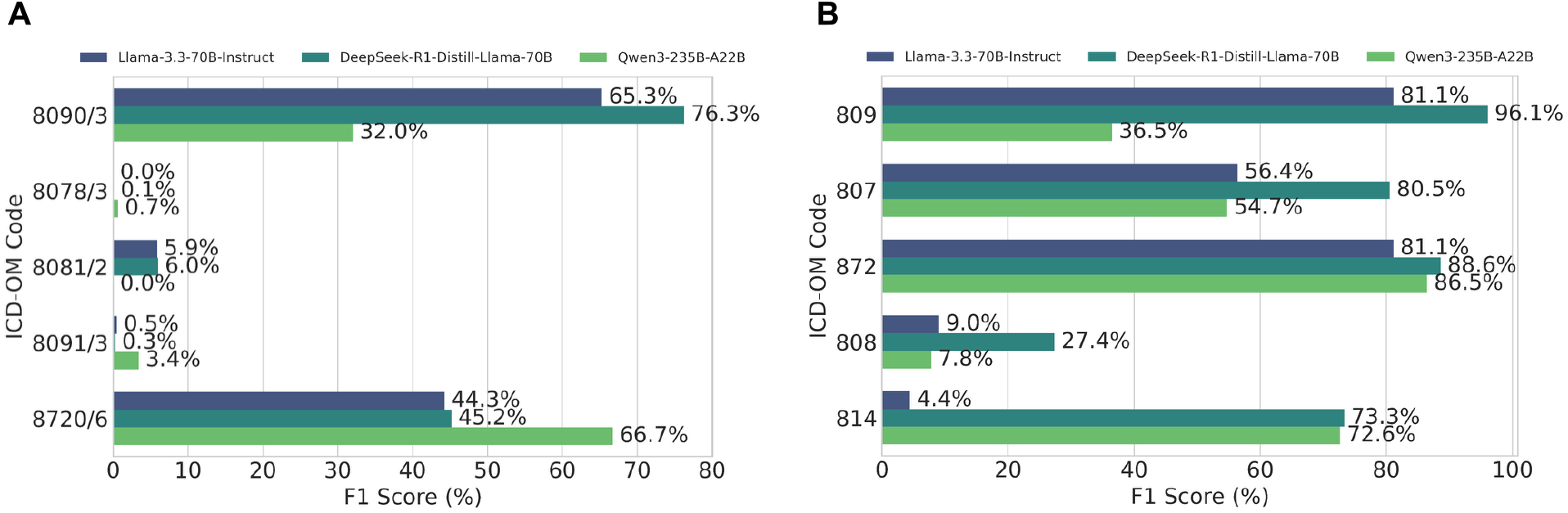
Performance in Relation to Support for ICD-OM Extraction Task. **A:** Exact Evaluation, **B:** First Three Character Evaluation Only the performance of the three best models for the five most frequent ICD-OM codes and groups are displayed. The order of the codes is descending, so the code displayed at the top of the plot is the most frequent code.

In contrast, the ICD-OM group evaluation showed greater variation in performance between models (**Fig. 6B**). For the most frequent class *809*, the F1-score ranged between 36.5% (*Qwen3-235B-A22B*) to 96.1% (*DeepSeek-R1-Distill-Llama-70B*). For other ICD-OM groups, such as *808*, the extraction showed lower F1-scores ranging from 7.8% (*Qwen3-235B-A22B*) to 27.4% (*DeepSeek-R1-Distill-Llama-70B*).

In addition to the ICD class, the text length of the pathology reports could potentially influence the model performance. To investigate this relationship, instance-based results for each report were calculated and averaged over the text length. The reports were categorized into eleven groups with text lengths ranging from 800 to 5200 (**Fig. 7**).

**Figure 7.**
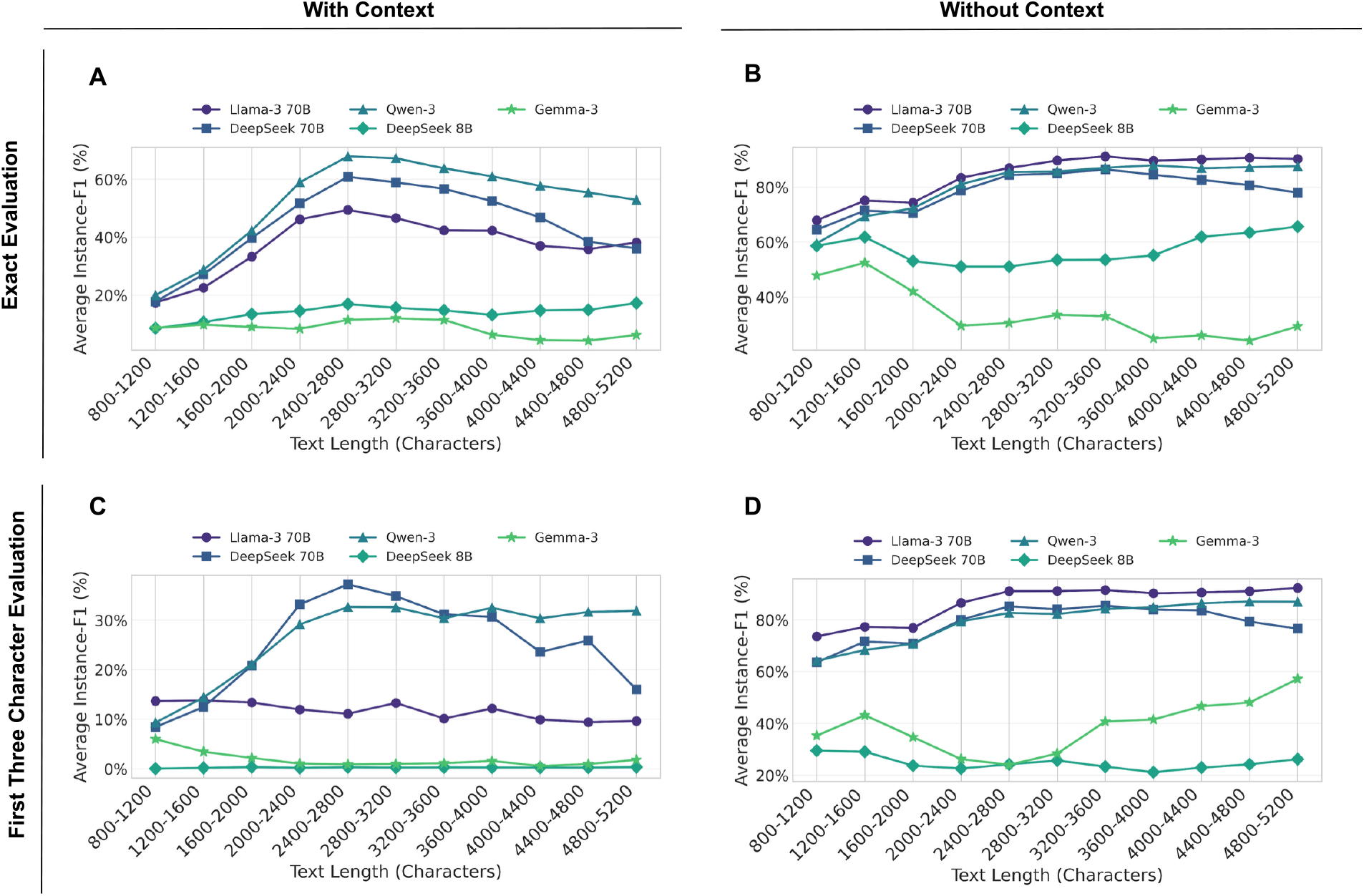
ICD-OT Performance in Relation to Text Length. **A:** Exact Evaluation with Context, **B:** Exact Evaluation without Context, **C:** First Three Character Evaluation with Context, **D:** First Three Character Evaluation without Context. The plots were created by calculating F1-scores for each report (instance-based). The text length was calculated by summing up the number of characters of each report and the lengths were then categorized into eleven groups.

For the exact ICD-OT task with context, shorter reports were classified with lower F1-scores on average (**Fig. 7A**). The highest performances were reached for reports with an average length between 2,000 and 2,400 characters. Longer reports in comparison were also classified with lower average scores. Models with reasoning capabilities (*DeepSeek-R1-Distill-Llama-70B* and *Qwen3-235B-A22B*) showed similar patterns for the first three character evaluation with context (**Fig. 7C**). In contrast, models lacking reasoning capabilities did not follow this pattern. Their performances showed that longer reports were classified with lower average scores.

Contrary results could be observed for the ICD-OT extraction task without context (**Fig. 7B**). Here, longer reports did not result in decreased performance They even scored best for *DeepSeek-R1-Distill-Llama-70B, Qwen3-235B-A22B*, and *DeepSeek-R1-Distill-Llama-70B* models. This pattern was consistent for both exact and first three character evaluation (**Fig. 7D**). Overall, the three best performing models achieved better performances on longer reports.

For the exact ICD-OM evaluation, model performance generally increased with longer reports (**Fig. 8A**). A similar trend was observed in the first three character ICD-OM evaluation (**Fig. 8B**). However, reports longer than 11,300 characters showed a greater variability in performance. Notably, the lowest scores were achieved for reports between 11,300 and 12,800 character length.

**Figure 8.**
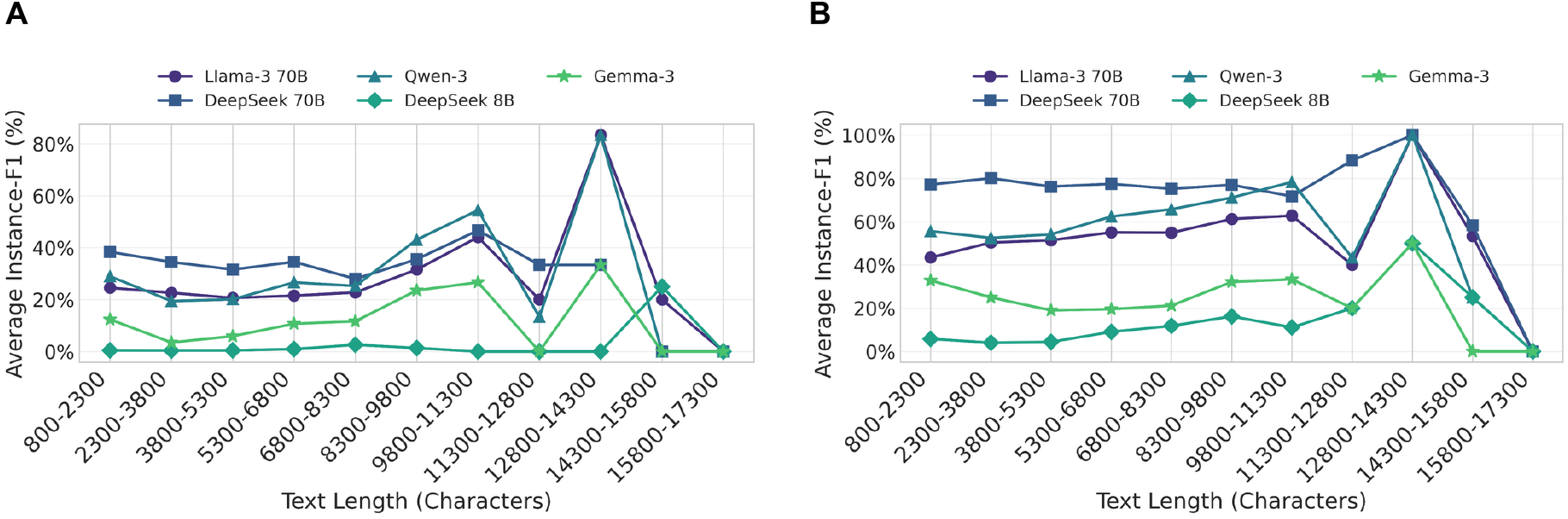
ICD-OM Performance in Relation to Text Length. **A:** Exact Evaluation, **B:** First Three Character Evaluation. The plots were created by calculating F1-scores for each report (instance-based). The text length was calculated by summing up the number of characters of each report and the lengths were then categorized into eleven groups.

## Discussion

Manual coding of pathology reports is a critical yet labor-intensive process in hospital information systems. As healthcare systems seek automation to improve efficiency and reduce errors, large language models (LLMs) have emerged as promising tools for extracting structured information from clinical texts. This study systematically evaluated the performance of several state-of-the-art open-source LLMs in predicting ICD-O-3 topography and morphology codes from real-world pathology reports. Our findings highlight that model performance is highly dependent on task type, input context, and evaluation criteria. While *Qwen3-235B-A22B* achieved the highest performance for exact and broad ICD-O topography code prediction when context about anatomical locations was provided in the prompt, *Llama-3*.*3-70B-Instruct* excelled in broader three-character classification when no additional context was provided, and *DeepSeek-R1-Distill-Llama-70B* demonstrated superior performance in morphology code extraction.

The substantial disparity between micro-average and macro-average performance metrics across all models reveals a limitation in clinical applicability, indicating that while the models perform well on frequently occurring codes, they struggle with rare conditions. This pattern was consistent across all evaluated models and both coding tasks. The performance improvement when evaluating only the first three characters suggests that current LLMs are better at identifying general anatomical categories than making fine-grained distinctions. This aligns with previous findings by Thrun et al.^14^ and Huang et al.^15^, who reported high accuracy rates (>90% and 89% respectively) for broader pathological classifications. Similarly, Soroush et al.^42^ observed that even larger proprietary LLMs, such as GPT-4 or Gemini, had difficulty with exact ICD codes, but often generated conceptually relevant outputs. The superior performance of larger models compared to smaller ones underscores the importance of model scale for capturing clinical nuances. However, the computational demands of large models present challenges for their deployment in clinical settings. Another finding from the evaluation was the dramatic performance degradation when anatomical context was removed from the prompts. The *Qwen3-235B-A22B* model experienced a 44% relative performance drop (from 71.6% to 40.4% micro-average F1-score), while *DeepSeek-R1-Distill-Llama-70B* showed a 33% relative decrease (from 64.3% to 42.8% micro-average F1-score) when the anatomical meaning of ICD codes was not explicitly provided. This suggests that current LLMs do not effectively infer anatomical relationships and topographical coding principles from medical text alone, but rather rely heavily on explicit contextual information for accurate code assignment. Similar improvements from contextual prompting have been reported in other medical NLP tasks^43,44^. However, in some cases, the additional context reduced performance in our study. For example, *Llama-3*.*3-70B-Instruct* performed better on broad topography codes when no anatomical detail was provided (87.2 compared to 84.6 to micro-average F1-score). This suggests that excessive or irrelevant information may overwhelm the model or hinder its ability to generalize.

Our study has limitations, primarily related to its retrospective, real-world design. The analysis is based on pathology reports and clinical codes from a single German institution, potentially limiting generalizability due to differences in documentation and coding practices. To reflect clinical reality, we used routine clinical codes as ground truth, which may contain manual coding errors. Additionally, the dataset showed a strong class imbalance, with common, primarily skin-related conditions, disproportionately represented. However, this distribution mirrors real-world diagnostic frequency and may explain the observed performance differences between conditions. Proprietary models such as ChatGPT-4o or Gemini may achieve better results but were excluded due to data privacy constraints, reflecting the practical realities of healthcare settings. Furthermore, excluding reports due to context token-length constraints may have biased the dataset against more complex cases.

This work provides a foundation for large-scale evaluations of self-hosted LLMs in real-world coding tasks. While exact code prediction, particularly for morphology, remains a challenge, the models showed encouraging performance in broader topography classification. These findings highlight the evolving medical reasoning capabilities of LLMs, although their performance is not yet sufficient for unsupervised use in routine clinical practice.

## Acknowledgments

J.Keyl is supported by a German Research Foundation (DFG)-funded clinician scientist program (FU 356/12-2). The work of Bahadir Eryilmaz and Mikel Bahn was funded by a PhD grant from the DFG Research Training Group 2535 Knowledge- and data-based personalization of medicine at the point of care (WisPerMed).

The data for this project was provided by the Smart Hospital Information Platform (SHIP), managed by the Data Integration Center at the University Medicine Essen. SHIP serves as a comprehensive digital health platform for integrating data from all major clinical subsystems using a holistic FHIR-based approach. It enables the purification, analysis, distribution, and visualization of clinical data.

## Data Availability Statement

The data that support the findings of this study are not openly available due to reasons of sensitivity. Requests regarding the dataset can be sent to and will be reviewed.

