## Supplementary Material for "Automated ICD-O-3 Coding of Real-World Pathology Reports Using Self-Hosted Large Language Models"

#### Table of Contents

Supplementary Figures
- Supplementary Figure 1. Prompt Example ICD-O T with Context.
- Supplementary Figure 2. Prompt Example ICD-O T without Context.
- Supplementary Figure 3. Prompt Example ICD-O M.
- Supplementary Figure 4. Example response from the Deepseek model.
- Supplementary Figure 5. Micro-Average Results of ICD-OT Code Evaluation.
- Supplementary Figure 6. Micro-Average Results of Exact and Three Position ICD-OM Code Matches.

Supplementary Tables
- Supplementary Table 1. Overview of evaluated large language models and their parameter sizes.
- Supplementary Table 2. Sampling parameters used for inference of LLMs.
- Supplementary Table 3. Complete Evaluation Results.

### Supplementary Figures

#### Supplementary Figure 1. Prompt Example ICD-O T with Context.

Prompt that was used to identify the anatomical location mentioned in the report. The report originally was documented in the hospital information system at the investigating site but anonymized for this figure to ensure data privacy. The prompt including the task description and report were originally written in German and translated to English for demonstration purposes.

**Instruction Sample:**

```
### Task: Determine the most appropriate ICD-O-3 classification topography
code for the anatomic location of the tumor from the following pathology
report. Select the best matching code from the following list and return
only the code with no additional explanation or description.
If no exact match is possible, enter the closest code.

### List of anatomical locations:

Lip (C00.), Outer upper lip (C00.0.), Outer upper lip (C00.0.), …

### Pathology Report:

John Doe, *14.06.1963 report date 23.09.2024

Clinical details:
Follicular lymphoma initial grade 1 2017, grade 3a 2022, currently nodal
progression.

Macroscopy:
Lymph node punch biopsy inguinal right 2x: two punch cylinders with a
length of 3 cm each.

Microscopy:
Microscopically, two lymph node punch cylinders with predominantly
follicles of centrocyte-like cells, centroblasts can only be recognised
occasionally.
Immunohistochemically, the follicular proliferates are strongly positive
for CD20, CD10 and BCL2.
Cyclin D12 is negative. The proliferation rate (Ki-67) is around 15%.
Only a few T cells are found with CD3.

Diagnosis:
```

Right inguinal lymph node punch biopsy:
Infiltration by a follicular lymphoma of the classic type (WHO 4th edition grade 1-2).
No evidence of transformation in the present punch biopsy.

##### Generated Answer:
C77.3

### Supplementary Figure 2. Prompt Example ICD-O T without Context.

Prompt that was used to identify the anatomical location mentioned in the report. The report originally was documented in the hospital information system at the investigating site but anonymized for this figure to ensure data privacy. The prompt including the task description and report were originally written in German and translated to English for demonstration purposes.

**Instruction Sample:**

```
### Task: Determine the most appropriate topography code of the ICD-O-3
classification for the anatomical location of the tumor from the following
pathology findings.
Return only the code without additional explanation or description.

### Pathology Report:

John Doe, *14.06.1963 report date 23.09.2024

Clinical details:
Follicular lymphoma initial grade 1 2017, grade 3a 2022, currently nodal
progression.

Macroscopy:
Lymph node punch biopsy inguinal right 2x: two punch cylinders with a
length of 3 cm each.

Microscopy:
Microscopically, two lymph node punch cylinders with predominantly
follicles of centrocyte-like cells, centroblasts can only be recognised
occasionally.
Immunohistochemically, the follicular proliferates are strongly positive
for CD20, CD10 and BCL2.
Cyclin D12 is negative. The proliferation rate (Ki-67) is around 15%.
Only a few T cells are found with CD3.

Diagnosis:
Right inguinal lymph node punch biopsy:
Infiltration by a follicular lymphoma of the classic type (WHO 4th
edition grade 1-2).
No evidence of transformation in the present punch biopsy.

### Generated Answer:
C77.3
```

### Supplementary Figure 3. Prompt Example ICD-O M.

Prompt that was used to identify the morphology of the tumor mentioned in the report. The report originally was documented in the hospital information system at the investigating site but anonymized for this figure to ensure data privacy. The prompt including the task description and report were originally written in German and translated to English for demonstration purposes.

**Instruction Sample:**

```
### Task: Determine the most appropriate morphology code of the ICD-O-3
classification from the following pathology findings.
Return only the five-digit morphology code without any additional
explanation or description.
Notes on morphology coding: The morphology code describes the cell type of
the neoplasm and its biological behavior. Morphological code numbers
consist of five-digit code numbers from the number range 8000/0 to 9989/3.
The first four digits indicate the histological type of the tumor. The
fifth digit after the slash (/) indicates the biological behavior: 0 for
benign (benign), 1 for unclear dignity (borderline/uncertain), 2 for in
situ (non-invasive, premalignant), 3 for in malignant (malignant,
invasive), 6 for secondary/metastatic and 9 for malignant (non-specific).

### Pathology Report:

John Doe, *14.06.1963 report date 23.09.2024

Clinical details:
Follicular lymphoma initial grade 1 2017, grade 3a 2022, currently nodal
progression.

Macroscopy:
Lymph node punch biopsy inguinal right 2x: two punch cylinders with a
length of 3 cm each.

Microscopy:
Microscopically, two lymph node punch cylinders with predominantly
follicles of centrocyte-like cells, centroblasts can only be recognised
occasionally.
Immunohistochemically, the follicular proliferates are strongly positive
for CD20, CD10 and BCL2.
Cyclin D12 is negative. The proliferation rate (Ki-67) is around 15%.
Only a few T cells are found with CD3.

Diagnosis:
Right inguinal lymph node punch biopsy:
```

```
Infiltration by a follicular lymphoma of the classic type (WHO 4th
edition grade 1-2).
No evidence of transformation in the present punch biopsy.

### Generated Answer:
8090/9
```

#### Supplementary Figure 4. Example response from the Deepseek model.

The model was tasked to identify the anatomical location mentioned in the report. Some of the words in the examples were originally written in German and translated into English for demonstration purposes.

**<think>**
Okay, I need to figure out which entry from the provided list corresponds to the anatomical description in the pathology report. Let's start by understanding the report.

The report mentions two findings, I and II. Both are skin-related. Finding I is a well-differentiated squamous cell carcinoma (G2) of the Bowen type, and Finding II is an actinic keratosis. The locations are specified as "I pectoral re./II US re. medial."

Looking at the list, I see entries related to the skin, specifically under the C44 category. The relevant entries are:

- C44.0: Lip skin without further details
- C44.1: Eyelid
- C44.2: Outer ear
- C44.3: Skin of other and unspecified parts of the face
- C44.4: Haired scalp and skin on the neck
- C44.5: Skin on the trunk
- C44.6: Skin of the upper extremities and shoulder
- C44.7: Skin of the lower extremity and hip
- C44.8: Skin, several sub-areas overlapping
- C44.9: Skin without further details

The locations "pectoral" and "US re. medial" refer to the chest area, specifically the medial (toward the midline) part of the right upper quadrant of the breast. Since the chest is part of the trunk, the most appropriate code would be C44.5: Skin on the trunk. This code covers the skin of the trunk, which includes the chest area.
**</think>**

Answer: **C44.5**

### Supplementary Figure 5. Micro-Average Results of ICD-OT Code Evaluation.

Displayed are results of the different models on the evaluation dataset consisting of 15,608 pairs of pathology reports and ICD-OT codes.

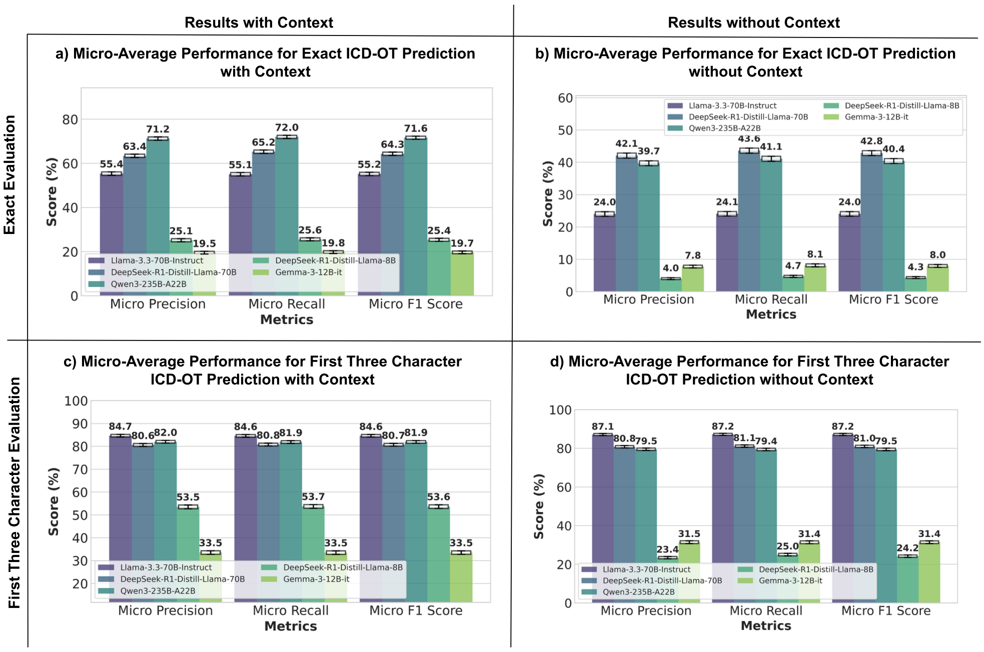


### Supplementary Figure 6. Micro-Average Results of Exact and Three Position ICD-OM Code Matches.

Displayed are results of the different models on the evaluation dataset consisting of 18,118 pairs of pathology reports and ICD-OM codes. This evaluation was calculated using exact and first three positions matches of ICD-code prediction.

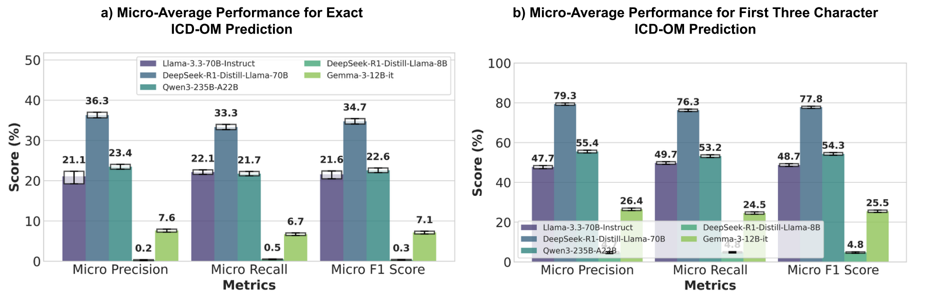

### Supplementary Tables

#### Supplementary Table 1. Overview of evaluated large language models and their parameter sizes.

The table includes both instruction-tuned base models (Llama-3.3, Gemma-3) and reasoning-enhanced variants (DeepSeek-R1-Distill, Qwen3-235B-A22B). Model sizes range from 8B to 235B parameters, with the Qwen3 model using a mixture-of-experts architecture that activates only 22B of its 235B parameters during inference.

| **LLM** | **Size** | **Architecture** |
|---|---|---|
| Llama-3.3-Instruct | 70B | Instruction-tuned |
| DeepSeek-R1-Distill-Llama | 8B/70B | Instruction-tuned |
| Qwen3-235B-A22B | 235B (22B activated) | Mixture-of-experts |
| Gemma-3-it | 12B | Instruction-tuned |

#### Supplementary Table 2. Sampling parameters used for inference of LLMs.

The following parameters were used for inference of the LLMs. For the *Qwen3* and *Deepseek* models, the recommended parameters by the creators of the model were used which can be found on the following web pages https://huggingface.co/Qwen/Qwen3-235B-A22B#best-practices and https://huggingface.co/deepseek-ai/DeepSeek-R1-Distill-Llama-70B#usage-recommendations.

| **Model** | **Top-P** | **Min-P** | **Top-K** | **Tempe-rature** | **Presence Penalty** |
|---|---|---|---|---|---|
| Qwen3 | 0.95 | 0 | 20 | 0.6 | 1.0 |
| Deepseek | 1 | - | - | 0.6 | 0 |
| Other models | 1 | - | - | 0 | 0 |

#### Supplementary Table 3. Complete Evaluation Results.

The table shows the performances of all evaluated models for each icd-code extraction task.

| ICD_Type | Model | Evaluation_Type | Micro_Precision | Micro_Recall | Micro_F1 | Macro-Precision | Macro-Recall | Macro-F1 |
|---|---|---|---|---|---|---|---|---|
| **icd-ot with context** | Llama-3 70B | Complete | 0.5537 [0.5458, 0.5615] | 0,5507 [0,543, 0,5583] | 0,5522 [0,5444, 0,5597] | 0,2418 [0,2382, 0,2756] | 0,3304 [0,3209, 0,3654] | 0,2533 [0,2505, 0,2858] |
| **iicd-ot with context** | Llama-3 70B | Three_Character | 0.8466 [0.8409, 0.8523] | 0,8456 [0,8399, 0,8512] | 0,8461 [0,8405, 0,8517] | 0,2255 [0,207, 0,2498] | 0,323 [0,2841, 0,3443] | 0,233 [0,2138, 0,2542] |
| **icd-ot with context** | Gemma-3 12B | Complete | 0.1954 [0.1893, 0.2014] | 0,1979 [0,1917, 0,204] | 0,1967 [0,1906, 0,2026] | 0,1156 [0,1156, 0,1364] | 0,1884 [0,1835, 0,2121] | 0,1221 [0,1215, 0,142] |
| **icd-ot with context** | Gemma-3 12B | Three_Character | 0.3353 [0.3276, 0.3430] | 0,3349 [0,3271, 0,3425] | 0,3351 [0,3274, 0,3428] | 0,1406 [0,1262, 0,1569] | 0,218 [0,1857, 0,2374] | 0,1401 [0,1318, 0,1563] |
| **icd-ot with context** | DeepSeek 70B | Complete | 0.6337 [0.6266, 0.6414] | 0,6524 [0,6451, 0,6602] | 0,6429 [0,6358, 0,6507] | 0,2462 [0,2446, 0,2799] | 0,3575 [0,3454, 0,3899] | 0,2637 [0,26, 0,2948] |
| **icd-ot with context** | DeepSeek 70B | Three_Character | 0.8058 [0.7996, 0.8120] | 0,8077 [0,8017, 0,8138] | 0,8067 [0,8007, 0,8129] | 0,2104 [0,1928, 0,2313] | 0,328 [0,2847, 0,344] | 0,2284 [0,2073, 0,2466] |
| **icd-ot with context** | DeepSeek 8B | Complete | 0.2514 [0.2446, 0.2583] | 0,2556 [0,249, 0,2623] | 0,2535 [0,247, 0,2603] | 0,1515 [0,1473, 0,1718] | 0,2472 [0,232, 0,2649] | 0,1631 [0,1567, 0,1809] |
| **icd-ot with context** | DeepSeek 8B | Three_Character | 0.5349 [0.5271, 0.5432] | 0,5369 [0,5291, 0,5451] | 0,5359 [0,5281, 0,5442] | 0,144 [0,1303, 0,1659] | 0,2249 [0,1924, 0,247] | 0,1406 [0,1313, 0,1598] |
| **icd-ot with context** | Qwen 3 235B | Complete | 0.7121 [0.7050, 0.7197] | 0,72 [0,713, 0,7273] | 0,716 [0,7092, 0,7233] | 0,2786 [0,274, 0,3117] | 0,391 [0,3817, 0,4288] | 0,2986 [0,2928, 0,3314] |
| **icd-ot with context** | Qwen 3 235B | Three_Character | 0.8197 [0.8140, 0.8258] | 0,8185 [0,8127, 0,8246] | 0,8191 [0,8133, 0,8252] | 0,2463 [0,2119, 0,266] | 0,3357 [0,296, 0,3575] | 0,2501 [0,2227, 0,2714] |
| **icd-ot without context** | Llama-3 70B | Complete | 0.2401 [0.2330, 0.2470] | 0,2408 [0,2339, 0,2476] | 0,2404 [0,2334, 0,2472] | 0,1828 [0,1837, 0,2147] | 0,2483 [0,2494, 0,2868] | 0,1812 [0,1821, 0,2126] |
| **icd-ot without context** | Llama-3 70B | Three_Character | 0.8712 [0.8657, 0.8765] | 0,8722 [0,8668, 0,8774] | 0,8717 [0,8663, 0,8769] | 0,2564 [0,2365, 0,2879] | 0,3011 [0,276, 0,3339] | 0,2382 [0,2257, 0,2674] |
| **icd-ot without context** | Gemma-3 12B | Complete | 0.0778 [0.0735, 0.082] | 0,0814 [0,0769, 0,0858] | 0,0795 [0,0752, 0,0838] | 0,0305 [0,0296, 0,0391] | 0,0402 [0,0402, 0,05] | 0,0259 [0,0258, 0,0333] |
| **icd-ot without context** | Gemma-3 12B | Three_Character | 0.3146 [0.3071, 0.3216] | 0,3142 [0,3067, 0,321] | 0,3144 [0,3069, 0,3213] | 0,0561 [0,055, 0,0678] | 0,067 [0,0595, 0,0802] | 0,0502 [0,0492, 0,0597] |
| **icd-ot without context** | DeepSeek 70B | Complete | 0.4206 [0.4128, 0.4284] | 0,4357 [0,4283, 0,4439] | 0,428 [0,4208, 0,4358] | 0,1899 [0,1878, 0,217] | 0,2933 [0,2819, 0,3178] | 0,2074 [0,2024, 0,232] |
| **icd-ot without context** | DeepSeek 70B | Three_Character | 0.8081 [0.8020, 0.8144] | 0,8115 [0,8056, 0,8175] | 0,8098 [0,8038, 0,8159] | 0,2085 [0,1914, 0,2444] | 0,2871 [0,2585, 0,3216] | 0,2038 [0,1914, 0,2351] |

| | | | | | | | | |
|---|---|---|---|---|---|---|---|---|
| **icd-ot without context** | DeepSeek 8B | Complete | 0.0401 [0.0374, 0.0430] | 0,0470 [0,0439, 0,0504] | 0,0433 [0,0404, 0,0464] | 0,0361 [0,0316, 0,039] | 0,0476 [0,0418, 0,0498] | 0,0375 [0,0325, 0,0396] |
| **icd-ot without context** | DeepSeek 8B | Three_Char acter | 0.2343 [0.2285, 0.2405] | 0,25 [0,2437, 0,2568] | 0,2419 [0,2359, 0,2483] | 0,011 [0,0113, 0,0147] | 0,0086 [0,0076, 0,0109] | 0,0076 [0,0078, 0,0099] |
| **icd-ot without context** | Qwen 3 235B | Complete | 0.3970 [0.3895, 0.4045] | 0,4111 [0,4033, 0,4186] | 0,4039 [0,3964, 0,4114] | 0,1489 [0,1502, 0,1731] | 0,2305 [0,2214, 0,2522] | 0,1611 [0,1608, 0,1842] |
| **icd-ot without context** | Qwen 3 235B | Three_Char acter | 0.7950 [0.7888, 0.8012] | 0,7942 [0,7877, 0,8004] | 0,7946 [0,7884, 0,8008] | 0,1935 [0,179, 0,2245] | 0,2298 [0,2151, 0,2626] | 0,1751 [0,1661, 0,2033] |
| **icd-om** | Llama-3 70B | Complete | 0.2106 [0.1922, 0.2235] | 0,2212 [0,2157, 0,227] | 0,2158 [0,2047, 0,2242] | 0,0229 [0,0193, 0,0496] | 0,0198 [0,0182, 0,0446] | 0,0154 [0,0138, 0,0351] |
| **icd-om** | Llama-3 70B | Three_Char acter | 0.4773 [0.4688, 0.4848] | 0,4974 [0,4908, 0,5042] | 0,4871 [0,4802, 0,4941] | 0,1511 [0,1396, 0,1922] | 0,1732 [0,1587, 0,2056] | 0,1264 [0,1182, 0,1544] |
| **icd-om** | Gemma-3 12B | Complete | 0.0761 [0.0723, 0.0797] | 0,067 [0,0637, 0,0702] | 0,0712 [0,0677, 0,0747] | 0,0062 [0,0052, 0,009] | 0,005 [0,0049, 0,0068] | 0,0037 [0,0036, 0,0049] |
| **icd-om** | Gemma-3 12B | Three_Char acter | 0.2644 [0.2584, 0.2707] | 0,2455 [0,2397, 0,2515] | 0,2546 [0,2488, 0,2607] | 0,0279 [0,0242, 0,0363] | 0,0237 [0,0228, 0,0293] | 0,0187 [0,0187, 0,0219] |
| **icd-om** | DeepSeek 70B | Complete | 0.3627 [0.3561, 0.3701] | 0,3332 [0,3269, 0,3398] | 0,3474 [0,3409, 0,3541] | 0,0813 [0,0822, 0,0986] | 0,0756 [0,077, 0,0917] | 0,065 [0,0669, 0,0789] |
| **icd-om** | DeepSeek 70B | Three_Char acter | 0.7932 [0.7878, 0.7993] | 0,7625 [0,7566, 0,7682] | 0,7776 [0,7721, 0,7831] | 0,3311 [0,3246, 0,3795] | 0,3607 [0,3485, 0,4055] | 0,3233 [0,3153, 0,3661] |
| **icd-om** | DeepSeek 8B | Complete | 0.0024 [0.0015, 0.0040] | 0,0047 [0,0038, 0,0057] | 0,0031 [0,0022, 0,0044] | 0,001 [0,0006, 0,0016] | 0,0001 [0,0001, 0,0003] | 0,0002 [0,0001, 0,0003] |
| **icd-om** | DeepSeek 8B | Three_Char acter | 0.0466 [0.0418, 0.0514] | 0,0484 [0,0454, 0,0516] | 0,0475 [0,0438, 0,051] | 0,0138 [0,0127, 0,0161] | 0,0105 [0,0095, 0,0128] | 0,0086 [0,0079, 0,0101] |
| **icd-om** | Qwen 3 235B | Complete | 0.2345 [0.2287, 0.2406] | 0,2175 [0,2122, 0,223] | 0,2256 [0,2202, 0,2315] | 0,0789 [0,0805, 0,095] | 0,0707 [0,0728, 0,0872] | 0,0597 [0,0615, 0,0732] |
| **icd-om** | Qwen 3 235B | Three_Char acter | 0.5545 [0.5481, 0.5617] | 0,5319 [0,5251, 0,5385] | 0,5429 [0,5365, 0,5498] | 0,3258 [0,3035, 0,36] | 0,3463 [0,3295, 0,3851] | 0,2984 [0,2825, 0,3306] |
